# Efficacy of multilevel exercise programmes with different intensities on development and metabolic homeostasis in overweight and obese children: study protocol for a randomized controlled trial

**DOI:** 10.1101/2023.06.09.23291210

**Authors:** Yimin Zhao, Chao Liu, Jiaxing Wang, Yangong Liu, Haiyi Yu, Tianqi Chang, Chenyu Fan, Wenxiu Wang, Yueying Li, Hu Wang, Yuzhou Xue, Wei Zhao, Chuan Ren, Ligang Cui, Xinheng Feng, He Huang, Dongwei Fan, Xiang Yao, Tongyan Han, Tao Huang, Ming Xu

## Abstract

**Background:** The prevalence of childhood obesity is rapidly increasing worldwide. Exercise has been suggested as the primary approach to manage body weight, but an effective exercise program tailored for school-age children is still lacking.

**Objective:** This trial aims to evaluate the efficacy of a novel multilevel (children, families, and schools)-coordinated exercise programmes with different intensities on adiposity and development among overweight and obese school-age children. It also aims to elucidate the complex interplays between the exercise interventions, multi-omic biomarkers, development, and metabolic homeostasis.

**Methods:** This is a multilevel –coordinated two-stage randomized double-blind controlled trial, comprising 366 overweight and obese children aged between 8 and 12 years from two elementary schools in Beijing, China. The novel multilevel exercise programs involve families and schools monitoring children’s exercise and diet via a specially designed smartphone application. The children will be randomly assigned to receive three school-based exercise programmes (exercise intervention stage), namely low-intensity exercise, moderate-intensity exercise, and intermittent vigorous exercise, for 12 weeks, followed by a 9-month post-intervention follow-up (follow-up stage). The primary outcomes of this trial are the changes in adiposity, development, and metabolic homeostasis during the two stages. The secondary outcomes include but are not restricted to genomics and changes in blood metabolomics, gut microbiota, and exercise performance.

**Discussion:** This trial has been approved by the ethics committee of Peking University Third Hospital (ethnic approval number 2021-283-08). The findings of this trial will help establish specialized multilevel coordinated school-based exercise programmes for overweight and obese school-age children in China and quantitatively and qualitatively evaluate the efficacy of the novel interventions.

**Trial registration:** This trial was prospectively registered at ClinicalTrials.gov with the identifier of NCT04984005 on July 21, 2021.

## Introduction

The prevalence of childhood obesity is rapidly increasing worldwide. According to recent data, the number of obese children and adolescents has increased from 11 million in 1975 to 124 million in 2016.^1 2^ This issue is becoming particularly prevalent in developing countries such as China, where the prevalence of childhood obesity and related non-communicable chronic diseases is rising rapidly.^3^ It is projected that by 2030, 6.64 million children under the age of seven, and 18.91 million children over the age of seven in China, will be obese.^4 5^ In Beijing, the prevalence of overweight and obesity was found to be 17.6% and 29.1% in boys and 17.6% and 18.0% in girls aged between 6 to 18 years, respectively.^5^

Childhood obesity has been linked to an increased risk of type 2 diabetes, heart disease, and chronic kidney disease in later life.^6–10^ While exercise intervention remains the primary approach for managing body weight, it may not achieve sustained desirable body weight reduction on its own for obese children who also have to manage academic stress.^11–14^ Indeed, a recent multicomponent lifestyle intervention successfully reduced adiposity and obesity prevalence among children.^4^ However, whether the involvement of schools and families could improve the effectiveness of exercise intervention among school-age obese children is still less studied.^15^ Notably, the parents and teachers may hold the opinions that strict exercise intervention may impede growth and development and academic performance of the children, thus reducing the effectiveness of exercise. In addition, it remains unknown whether a more intense exercise programme would bring about additional health benefits in terms of development and metabolic homeostasis beyond body weight during puberty for obese children.^1 14^

In this study, we aim to compare the effects of multilevel-coordinated exercise programmes with different intensities on adiposity, development, and metabolic homeostasis, as well as related mechanisms. Specifically, we aim to determine which exercise programme is more effective in reducing body weight and improving development and metabolic homeostasis among overweight and obese children, as well as identifying the factors that mediate exercise-induced changes in these areas. Additionally, we aim to establish a regionally representative school-family-hospital evaluation model for exercise intervention.

## Method and Analysis

### Trial design

This is a multilevel coordinated (children, families, and schools) two-stage 12-month three-arm randomized double-blind controlled trial.

### Methods: Participants, interventions, and outcomes

This protocol is reported following the guidelines of the Standard Protocol Items: Recommendations for Interventional Trials (SPIRIT) 2013 Statement.^16^

### Study setting

This trial will be conducted in two government-funded elementary schools in Beijing, the capital city of China. The steering committee of this study included paediatricians, physicians, epidemiologists, and statisticians from Peking University Third Hospital and School of Public Health at Peking University. All study procedures will be performed in accordance with the Declaration of Helsinki.

### Eligibility criteria

Eligible participants must meet the following inclusion criteria:

1) overweight or obese (see **Table 1** for age– and sex-specific BMI cutoff points);
2) 8 to 12 years old;
3) did not participate in any weight loss programmes, including medication, exercise, and lifestyle modification, within the last three months before baseline assessment;
4) provide written informed consent by both themselves and their statutory guardians.

**Table 1.** Age– and sex-specific BMI cutoff points of childhood overweight and obesity.^22^

|  | Boys |  | Girls |  |
| --- | --- | --- | --- | --- |
| Age, years | Overweight | Obesity | Overweight | Obesity |
| 8.0 - < 8.5 | 17.8 - < 19.7 | ≥ 19.7 | 17.6 - < 19.4 | ≥ 19.4 |
| 8.5 - < 9.0 | 18.1 - < 20.3 | ≥ 20.3 | 18.1 - < 19.9 | ≥ 19.9 |
| 9.0 - < 9.5 | 18.5 - < 20.8 | ≥ 20.8 | 18.5 - < 20.4 | ≥ 20.4 |
| 9.5 - < 10.0 | 18.9 - < 21.4 | ≥ 21.4 | 19.0 - < 21.0 | ≥ 21.0 |
| 10.0 - < 10.5 | 19.2 - < 21.9 | ≥ 21.9 | 19.5 - < 21.5 | ≥ 21.5 |
| 10.5 - < 11.0 | 19.6 - < 22.5 | ≥ 22.5 | 20.0 - < 22.1 | ≥ 22.1 |
| 11.5 - 12.0 | 19.9 - < 23.0 | ≥ 23.0 | 20.5 - < 22.7 | ≥ 22.7 |

Participants will be excluded if they have any of the following conditions:

1) have any severe chronic disease, including asthma, cardiovascular disease, and cancer.
2) have drug-induced secondary obesity;
3) have Cushing syndrome, Prader-Willi syndrome, or other medical conditions that may lead to obesity;
4) have intellectual, mental, or behavioural disorders;
5) feel difficulty communicating with physical education (PE) teachers, paediatricians, and physicians;
6) have any other conditions that may prevent exercising safely, which will be assessed by paediatricians and physicians.

### Who will take informed consent?

A group of research staff, including paediatricians and physicians, will collect written informed consent from both children and their statutory guardians before evaluating the eligibility of potential participants. During the consent process, potentially eligible participants and their parents will be notified that this trial will require the collection of fasting blood, urine, and faeces samples.

## Interventions

### Explanation for the choice of comparators

Eligible participants will be randomly assigned into three groups receiving low-intensity exercise, moderate-intensity exercise, and intermittent vigorous exercise, respectively. The low-intensity exercise is chosen as the comparator because exercise intervention, even at a low intensity, has been proven effective in improving energy expenditure and cardiometabolic health among children and adolescents. The novel intervention programmes emphazie the involvement of family and school to assist children’s exercise and food intake using a specifically designed smartphone application. The health educators, the pediatrician in the school hospital, will be trained by the project staff before the intervention. During the intervention, the health educator will deliver two health education sessions, including exercise and healthy eating, to children every month. Family members and school PE teachers at each site will be separately invited to attend the health education sessions two times per month, where the project staff shared the information on children’s exercise performance and strengthened the family and school involvement to assist children’s exercise.

### Intervention description

The levels of intensity will be determined according to the percentages of heart rate reserve (HRR) during exercise, which is calculated by deducting the resting heart rate (HR_resting_) from the maximum heart rate (HR_max_).^17 18^ The HR_resting_ and the HR_max_ will be evaluated on a COSMED Quark PFT pulmonary function testing system in Peking University Third Hospital at baseline.

All participants will engage in a school-based exercise programme, instructed by PE teachers, at the frequency of 30 minutes/time × two times/day × three days/week for 12 weeks. Each 30-minute exercise will comprise a 7-minute warm-up session, a 16-minute exercise session, and a 7-minute cool-down session. In the warm-up and cool-down sessions, all groups of participants will do walking and stretching exercises, including arm circles, overhead arm reaches, and hip rotations, to reach a target heart rate of around 0.3 × HRR + HR_resting_. During the 16-minute exercise session:

1) for the low-intensity exercise group, the participants will be instructed to take 800-meter quick walking and high knees to reach a target heart rate between 0.3 × HRR + HR_resting_ and 0.4 × HRR + HR_resting_;
2) for the moderate-intensity exercise group, the participants will be instructed to take 30-meter S-shaped quick walking, 30-meter S-shaped running, and 800-meter alternating between running and walking to reach a target heart rate between 0.4 × HRR + HR_resting_ and 0.69 × HRR + HR_resting_;
3) for the intermittent vigorous exercise group, the participants will be instructed to take various 30-second jumping and quick running activities to reach a target heart rate between 0.7× HRR + HR_resting_ and 0.85 × HRR + HR_resting_, divided by a 60-second relaxation above 0.6× HRR + HR_resting._

In the 2-week run-in period prior to intervention period, all participants will receive the same low-intensity exercise intervention as described above.

### Criteria for discontinuing or modifying allocated interventions

We allow participants to discontinue the exercise intervention at any time for any reason. The research staff will document the reasons for dropping out if provided by the participants. We will not recruit new participants to fill the vacancy after the implementation of the exercise intervention.

### Strategies to improve adherence to interventions

Participants will be instructed to wear a calibrated smartwatch (Huawei Band 6) during the run-in and intervention stages, which is provided by Huawei Technologies for free. During each 30-minute exercise session, the smartwatch will return 30 reads of real-time heart rate at a 1-minute interval. Adherence to the exercise programme is assessed by counting the real-time heart rate reads that are within the target range for each participant. We assume that all 16 reads collected during the 16-minute exercise session are located within the target heart rate zone in each group. In addition, we will reward participants who satisfactorily complete the 12-week exercise intervention with a gift, including a pen and pencil box.

### Relevant concomitant care permitted or prohibited during the trial

During the 2-week run-in period and 12-week exercise intervention period, all participants will be instructed to maintain their usual physical activity and diet using the specifically designed smartphone application. We will monitor their diet compositions via a three-day dietary recall, and the nutrient intake will be analyzed according to China Food Composition Tables.^19^ We will assess their physical activity, excluding exercise intervention, via the Huawei band 6. These two questionnaires will be performed by trained research staff face-to-face. In addition, participants and their parents will receive notifications via a mobile phone application, which will notify the participants to maintain their usual diet and physical activity.

### Provisions for post-trial care

We will implement a 9-month post-intervention follow-up to observe the dynamic changes in adiposity among the participants.

## Outcomes

### Primary outcomes

The primary outcomes of this trial are the changes in adiposity and biomarkers of development and homeostasis, including anthropometric measurements, sexual hormones, body compositions, cardiopulmonary functions, and biochemical measurements.

### Secondary outcomes

Secondary outcomes include changes in:

1. Exercise performance;
2. Hepatic hemodynamic index;
3. Bone mineral density;
4. Inflammatory biomarkers;
5. Thyroid function;
6. Oxidative stress biomarkers;
7. Oral and gut microbiota;
8. Blood and faecal metabolomics;
9. Flow-mediated dilation
10. Attention performance;
11. Sleep quality;
12. Diets and nutrients;
13. Whole genome sequencing;
14. Viromics;
15. Proteomics;
16. Volatile organic compounds in breath;

### Participant timeline

**Figure 1** shows the schedule of enrollment, allocation, interventions, and assessments.

**Figure 1.**
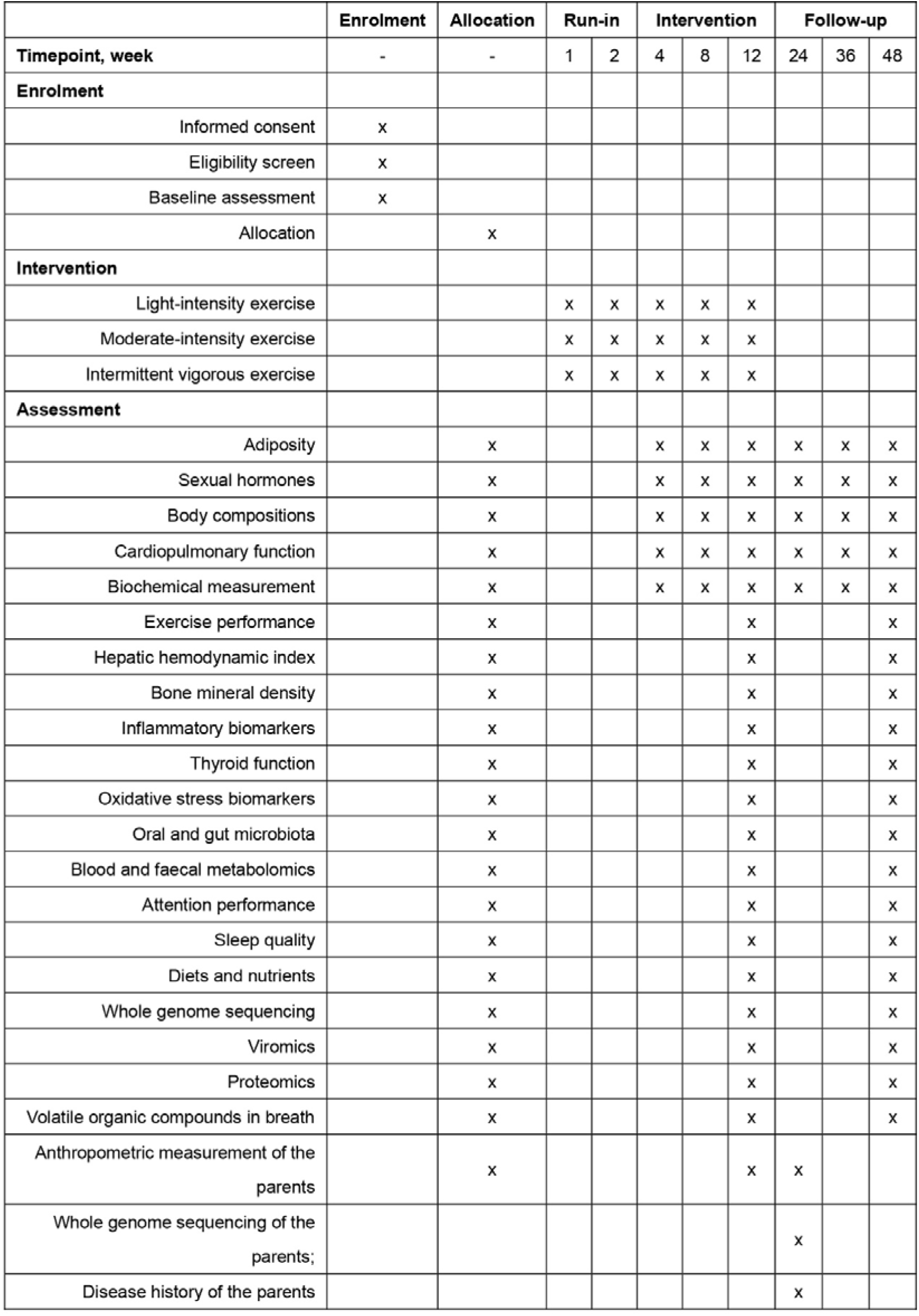
Schedule of enrollment, allocation, interventions, and assessments.

### Sample size

We estimated the sample size using the following equation:

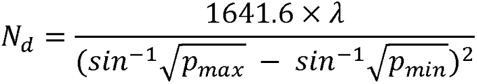

where N_d_ is the minimum sample size, P_max_ and P_min_ are the highest and lowest cure rates, and λ depends on α, β, and degree of freedom. In this trial, α is 0.05, β is 0.1, and the degree of freedom is 2. Thereby, λ is 12.65. We assumed that 65%, 50%, and 40% of overweight and obese participants that receive low-intensity exercise, moderate-intensity exercise, and intermittent vigorous exercise would successfully lower their body weight, achieving a healthy BMI, respectively. A minimum sample size of 122 is required for each group accounting for a 10% drop-out rate. A total of 366 children will be included in this trial.

### Recruitment

From January to March 2022, we included two elementary schools in Beijing by inviting the school principals to participate in this trial. The two schools agreed with the study protocol. We will recruit eligible participants, aged between 8 and 12 years, from seven school campuses. All students and their parents (or their statutory guardians) will receive an advertisement for this trial from the principal and their PE teachers. Potential eligible participants who provide written informed consent by themselves and statutory guardians will be asked to complete a nurse-administrated questionnaire concerning their health-related information and undergo a physical examination according to standard operating procedure.

## Assignment of interventions: allocation

### Sequence generation

We will perform sex– and BMI-stratified randomization via computer-generated random numbers.

### Concealment mechanism

The participants will be randomized using a computer-generated random sequence, which will be sealed in an envelope until the end of this trial.

### Implementation

An independent statistician from the School of Public Health, Peking University, who will not participate in recruitment, allocation, baseline assessment, and intervention, will conduct the randomization.

## Assignment of interventions: Blinding

### Who will be blinded

All participants, their parents, the PE teachers, the paediatricians, the physicians, and the statisticians will be blinded to the allocation.

### Procedure for unblinding if needed

Not applicable to this trial.

## Data collection and management

### Plans for assessment and collection of outcomes

All data will be collected and processed according to standard operating procedures. The demographic information of participants will be collected at baseline. Trained nurses will conduct the physical examination under the supervision of paediatricians. The biochemical measurements will be conducted at the clinical laboratory of Peking University Third Hospital. The oral and gut microbiota compositions will be analyzed using the QIIME2 pipeline (https://qiime2.org). The metabolomics analysis will be conducted on a liquid chromatography-electrospray ionization tandem mass spectrometry system (QTRAP 6500+ system).^20, 21^ Diet will be evaluated via a 3-day diet recall at baseline and at the end of intervention period. Meanwhile, the participants will be instructed to take photo of meals to facilitate nutrient intake analysis. Nutrients intake will be analzyed according to the China Food Composition Tables by two independent investigators.^19^

### Plans to promote participant retention and complete follow-up

No other strategy is planned to promote participant retention and complete follow-up.

## Data management

A double entry and validation approach is planned for data management. The personal information and medical records of each participant will be kept in an individual folder and stored in a secure storage area at the School of Public Health, Peking University. We will return the results of the physical examination and laboratory biochemical measurement to participants at the end of the exercise intervention/follow-up.

## Confidentiality

The personal information and medical records of participants will be treated as strictly confidential. The research team will ensure the privacy and confidentiality of the personal information and medical records of participants and store all data in an encrypted database with all identifiers removed.

### Plans for collection, laboratory evaluation and storage of biological specimens for genetic or molecular analysis in this trial/future use

At baseline and the end of exercise intervention, 6 mL overnight fasting venous blood will be collected by trained nurses from Peking University Third Hospital under the supervision of a paediatrician. The participants will also provide urine and faeces samples. All biospecimens will be immediately delivered to the clinical laboratory of Peking University Third Hospital and stored at –80℃ until analysis.

## Statistical methods

### Statistical methods for primary and secondary outcomes

Statistical analysis will be performed using Stata/MP version 17.0 by statisticians from Peking University Health Science Center, which will not participate in participant enrollment, allocation, and exercise intervention. The intention-to-treatment analysis will be used, which includes all participants who complete the baseline assessment and is subject to randomization and exercise intervention. Between-group differences in continuous and categorical variables at baseline will be analyzed using one-way analysis of variance and chi-square test, respectively. Within-group changes from baseline to the end of the intervention will be analyzed using paired t test for each group. Between-group differences of changes in the continuous variables will be analyzed using one-way ANOVA, one-way analysis of covariance, or Mann-Whitney rank sum test when appropriate. Correlations of changes in continuous variables will be analyzed using Pearson’s or Spearman’s correlation analysis when appropriate. Statistical significance is set at α = 0.05.

### Interim analyses

No interim analysis will be performed due to the short duration of this trial.

### Methods for additional analyses (e.g. subgroup analyses)

In the sensitivity analysis, per protocol analysis is planned for participants that satisfactorily complete both the baseline assessment and the two-stage intervention.

### Methods in analysis to handle protocol non-adherence and any statistical methods to handle missing data

Missing covariates will be estimated using the multiple imputation method.

### Plans to give access to the full protocol, participant level-data and statistical code

Access to the full study protocol, participant-level data, and statical codes will be granted for authorized research staff and will not be publicly available.

## Oversight and monitoring

### Composition of the coordinating centre and trial steering committee

The principal investigator (Prof. Ming Xu in the author list) has full responsibility for the design and conduct of the trial. The steering committee is responsible for agreeing on the final protocol and reviewing the progress of the trial and potential changes to the protocol.

### Composition of the data monitoring committee, its role and reporting structure

The data monitoring committee is composed of pedestrians and statisticians and is responsible for assessing adherence to exercise intervention of each participant, monitoring and reporting any adverse events during exercise intervention.

### Adverse event reporting and harms

The research team is responsible for reporting any adverse events. During exercise intervention days, a field team comprised of a paediatrician (or a physician) and a research staff from the site management organization will be sent to each school to deal with potential adverse events. The PE teachers and other elementary school teachers will assist in addressing potential anxiety and mood issues during exercise intervention. All severe adverse events will be reported to the ethnic committee of Peking University Third Hospital, which will decide whether to continue or terminate this trial.

### Frequency and plans for auditing trial conduct

The principal and lead investigators will meet biweekly to review the progress of the trial and audit the trial conduct.

### Plans for communicating important protocol amendments to relevant parties (e.g. trial participants, ethical committees)

Any amendments to the study protocol must receive approval from the Ethics Committee of Peking University Third Hospital. All participants, their parents, PE teachers, and research staff will be promptly informed of any changes made to the study protocol. In case of a potential COVID-19 outbreak in the school, the intervention will be suspended and resumed in compliance with the school and government policies.

## Dissemination plans

The results of this trial will be presented at domestic and international conferences and published in peer-reviewed academic journals. Progress and final reports will be submitted to the funding agency and regulatory bodies.

## Conclusions

The present trial will provide a novel multilevel-coordinated exercise intervention targeting obese and overweight children. This study aims to yield quantitative and qualitative results which will shed light on the effectiveness of multi-level exercises with various intensities in promoting cardiometabolic fitness and reducing adiposity in overweight and obese school-aged children. Moreover, this study will utilize a multi-omic approach and systematic epidemiological design to investigate the possible molecular mechanisms of the exercise intervention in modulating adiposity at multiple levels. The outcomes of this trial will offer clinical evidence for establishing bodyweight management guidelines for Chinese schools.

## Trial status

This trial has been registered at ClinicalTrials.gov with the identifier NCT04984005.

## Data Availability

All data produced in the present study are available upon reasonable request to the authors.

## Acknowledgements

Our special thanks go to the children, parents and physical education teachers who are participating in this study. We also acknowledge the ethics committee of Peking University Third Hospital. We thank Huawei Technologies Co., Ltd. for donating the smartwatch and the technical support of the Huawei Research platform to this study. We also thank Beijing BOE Health Technology Co., Ltd. for providing the equipment for the physical examination.

## Author contributions

YZ, CL, TH, and MX contributed to conception and design of the study. JW, YL, and HY organized the database. YZ, CL, TC, CF, WW, YL, HW, YX, WZ, CR LC, XF, DF, HH, XY, and TH performed the study. YZ and CL wrote the first draft of the manuscript. All authors contributed to manuscript revision, read, and approved the submitted version.

## Availability of data and materials

Study data will be available from the principal investigator upon reasonable request and will not be made publicly available.

## Ethics approval and consent to participate

This study has been approved by the ethics committee of Peking University Third Hospital (ethnic approval number 2021-283-08).

## Consent for publication

Not applicable.

